# Eight decades of follow-up link life course exposures to proteomic organ ageing and longevity

**DOI:** 10.1101/2025.09.07.25335188

**Authors:** James W Groves, Veronica Augustina Bot, Daisy Yi Ding, Jennifer Nicholas, Amelia Farinas, Hamilton See-Oh, Sarah-Naomi James, Andrew Wong, Dylan M Williams, Josh King-Robson, Nishi Chaturvedi, Marcus Richards, Tony Wyss-Coray, Jonathan M Schott

**Affiliations:** Dementia Research Centre, UCL Queen Square Institute of Neurology, London, UK; Wu Tsai Neurosciences Institute, Stanford University, Stanford, CA, USA; The Phil and Penny Knight Initiative for Brain Resilience, Stanford University, Stanford, CA, USA; Department of Medical Statistics, London School of Hygiene and Tropical Medicine, London, UK; MRC Unit for Lifelong Health and Ageing, University College London, London, UK; Division of Psychiatry, University College London, London, United Kingdom; Department of Neurology & Neurological Sciences, Stanford University School of Medicine, Palo Alto, California, USA; UK Dementia Research Institute at UCL, London, UK

## Abstract

The pace of organ ageing varies substantially between individuals, yet drivers of variability remain poorly understood. This gap is critical, given only 20-30% of longevity is genetically inherited^1,2^ and age-related diseases are leading causes of morbidity and mortality^3^. Proteomic clocks allow organ ageing to be estimated from blood sampling^4^, facilitating study of how life course exposures shape biological ageing heterogeneity. Here, we leverage the unique design of the MRC National Survey of Health and Development (NSHD), the world’s oldest continuously followed birth cohort, to track 1,803 individuals across eight decades since birth in 1946. At mean age 63.2 years, we estimated proteomic ageing in seven organs. Despite near identical chronological ages, participants’ proteomes revealed biological ageing disparities spanning decades. Extreme ageing in multiple organs was a strong prognostic indicator for all-cause mortality over the following 15 years (HR=6.62 for ≥ 4 extremely aged organs). Adversity and being overweight in adolescence associated with accelerated ageing decades later in life. Completing secondary school education and maintaining physical activity linked to relative biological youth. Mediation analyses indicated liver, kidney and immune ageing linked life course exposures to mortality. Across 10,776 plasma protein targets, we identified 143 predictors of longevity, including MED9, strongly linked to diverse socio-behavioural exposures. These findings provide unique insights into which factors are likely to shape how we age, when in life they may be influential, and how biological effects emerge, informing healthy ageing promotion.

## MAIN TEXT

Ageing is a biological process driving susceptibility to chronic disease and mortality^3^. Although all humans undergo ageing, there is significant inter-individual variation in its speed and pattern of progression^5,6^. Biological tools quantifying this variation facilitate investigation into how life circumstances and behaviour alter ageing trajectories, with potential insights for personalised health promotion^7^.

Dysfunction in protein homeostasis is a hallmark of ageing^8^. Recently developed proteomic age estimators (“proteomic ageing clocks”) leverage age-related patterns in circulating proteins to quantify the amount of ageing an individual is estimated to have undergone on a molecular level: their ‘biological age’, which may deviate markedly from chronological age^9–12^. This approach has been extended to develop organ-specific age estimators, allowing the molecular ages of different organs to be compared within individuals^4^.

However, it remains largely unknown which modifiable factors influence molecular organ ageing and when in the life course their effects are most pronounced. This is a critical gap, given that only 20-30% of longevity is thought to be genetically inherited^1,2^. Birth cohort studies, which prospectively track comparable individuals across the life course, provide a unique opportunity to explore the shaping influence of diverse risk factors, capture cumulative exposure effects, identify whether certain life periods are particularly influential, and implicate specific biological pathways^13,14^.

In pursuit of this, we undertook a life course investigation of 1,803 members of the MRC National Survey of Health & Development (NSHD): the world’s longest continuously studied birth cohort^15^. By tracking identically aged participants, geographically representative of the British population, across eight decades, we: (i) investigated variability in proteomic signatures of organ ageing between closely comparable individuals and assessed its prognostic value for longevity; (ii) evaluated whether life course exposures were linked to organ ageing variation and (iii) characterised the biological interfaces of longevity and socio-behavioural exposures through an analysis of 10,776 protein targets **(Fig. 1e)**.

**Figure 1.**
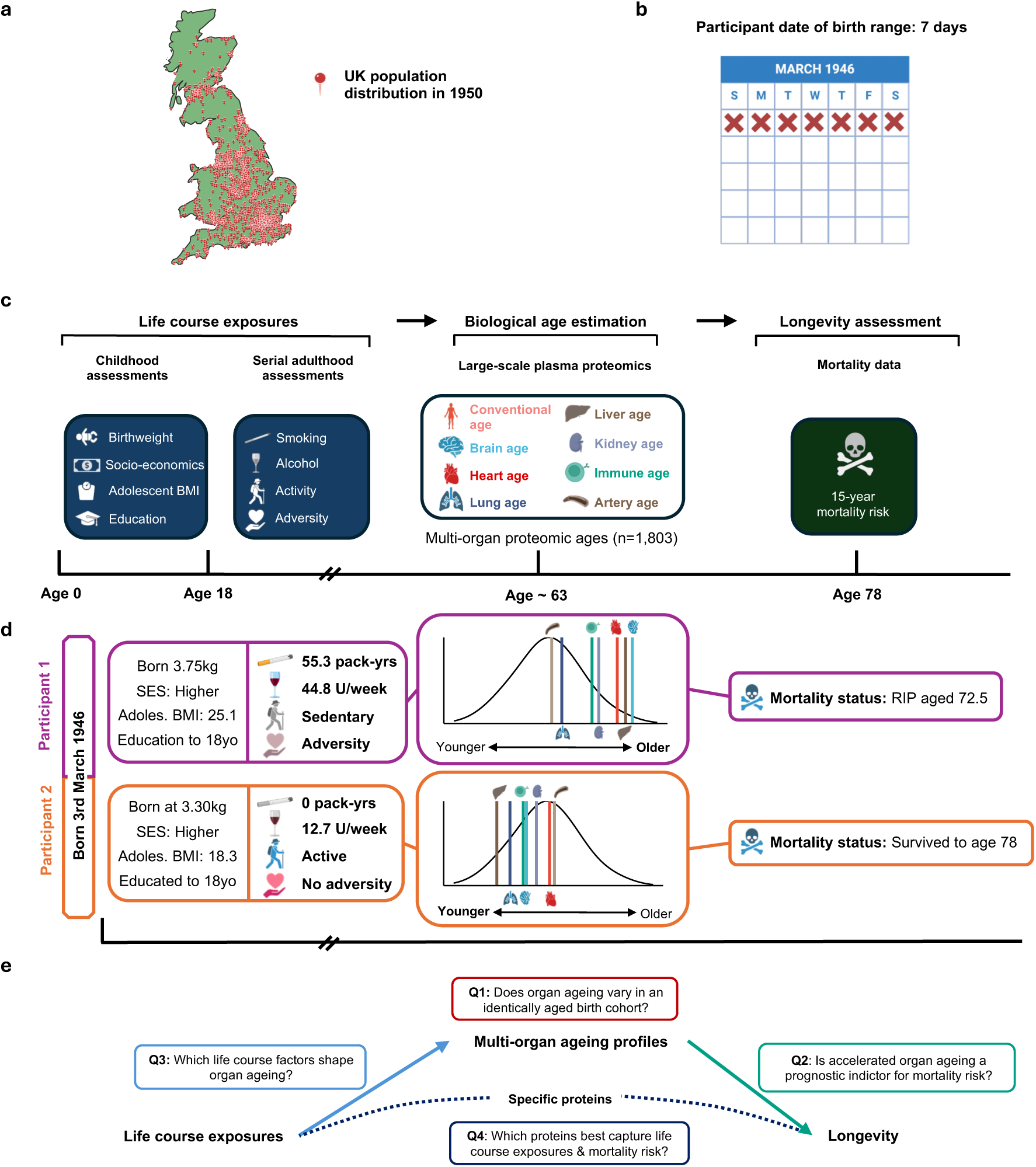
Study design. **a**, Participants were 1,803 individuals from the National Survey of Health and Development (NSHD): a sample geographically representative of mainland Britain in 1946. Illustrative map of mainland Britain with pins placed according to 1950 population density from the History Database of the Global Environment (HYDE). All participants were born in a single week in March 1946 (**b**). **c,** Timeline of measurements taken from participants’ birth to old age. We assessed life course exposures, proteomic multi-organ ageing phenotypes and longevity. **d**, Illustrative examples highlighting the data collected throughout the participants’ life course. These hypothetical individuals with differing risk factor profiles, organ ageing estimates and longevity showcase the heterogeneity observed in life circumstances, health behaviours and outcomes. **e**, Schematic of the questions this study aims to address using its life course approach. Figure elements created with BioRender.com.

### A life course design capturing exposures, proteomic organ ageing and longevity

We included 1,803 individuals from NSHD **(Extended Data Fig. 1; Supplementary Table 1)**, a geographically representative sample of the population of mainland Britain **(Fig. 1a)** born in a single week in March 1946 **(Fig. 1b)**. Plasma was sampled in late midlife (63.2 ± 1.1 yrs) with the 11k SomaScan (v5.0) assay^16^ to take 10,776 human protein measurements, ∼3,000 of which, to our knowledge, were previously unevaluated in population studies of ageing or longevity.

Proteins covered by the previous 5k SomaScan assay (v4.0) were used to generate biological age estimates for each participant, using published algorithms **(Fig. 1c; Supplementary Table 2)** ^4^. The ‘Conventional age’, based on the abundance of all measurable age-related proteins, represents an over-arching biological age estimate for each individual, largely determined by non-organ-specific proteins **(Supplementary Table 2)**^4^. Organ age estimates are based on abundance of proteins enriched for a specific organ^4^. For example, 201 protein targets enriched in brain tissue contribute to the brain age estimate, while 103 liver-enriched proteins contribute to the liver age estimate, where no protein is assigned to more than a single organ. Focussing on organs with strong links to mortality, we studied organ ageing in the brain, heart, lungs, liver, kidneys, immune system, and arteries **(Fig. 1c)**^17^.

We measured eight socio-behavioural factors: four in childhood and four serially across adulthood **(Fig. 1c)**. Since measures of growth and development may associate with epigenetic ageing^18,19^, we studied two anthropometric traits bookending childhood: low birthweight and overweight in adolescence. Social disadvantage was recently connected to accelerated proteomic ageing^20^, so we explored if higher childhood socio-economic status and educational attainment might drive favourable organ-specific trajectories.

A major advantage of studying a birth cohort is access to prospectively collected data from birth, avoiding potential inaccuracies of retrospective reports^21^. We evaluated serially acquired prospective data on smoking, alcohol and physical activity throughout adult life. We utilised a published adversity score^22^, which captured financial and psychosocial stressors across the life course, including unemployment, divorce and social isolation.

Finally, mortality was tracked through data-linkage to health care records up to age 78.3 years (mean follow-up from proteomic sampling 15.1 ± 1.2 yrs) **(Fig. 1c)**. Thus, participants were continuously followed from birth to near the mean projected lifespan in 1946^23^ **(Fig. 1d)**. We hypothesised that maintaining a favourable life course exposure profile would reduce risk for accelerated ageing by late midlife, lessening chance of early death **(Fig. 1e)**.

### Substantial organ ageing divergences strongly stratify mortality risk

Proteomic ageing profiles were expressed as age gaps. These reflect disparity between a participant’s biological age and their actual chronological age, which, as all cohort members are born in the same week, is essentially identical. *Positive* values indicate the number of years biologically *older* a participant appeared relative to their chronological age, whilst *negative* values reflected a biologically *younger* phenotype^4^.

Distributions of Conventional and organ-specific ageing **(Fig. 2a)** revealed marked variation in biological ageing between participants. For brain ageing, despite mean age gap being very close to true chronological age (-0.3 yrs), over forty years separated the biologically youngest and oldest participants (range: -19.8, 25.7 yrs), with more than thirty years of divergence seen in liver (mean: -1.3 ± 4.2 yrs; range: -15.6, 21.5 yrs) and immune (mean: 0.13 ± 3.7 yrs, range: -12.9, 22.7 yrs) ageing **(Supplementary Table 3)**. 44.1% of participants had extreme ageing in at least one organ (defined as the top decile of each age gap distribution)^12^, with 16.0% having two or more organs affected. Ageing across different organs was heterogeneous and not highly correlated (Pearson’s *r* < 0.4 for all organ combinations; **Supplementary Table 4**). Brain, immune and liver age gaps were the most closely related **(Extended Data Fig. 2a),** and these organs most likely to undergo extreme ageing concurrently **(Extended Data Fig. 2b; Supplementary Table 5)**, potentially reflecting their physiological interdependence.

**Figure 2.**
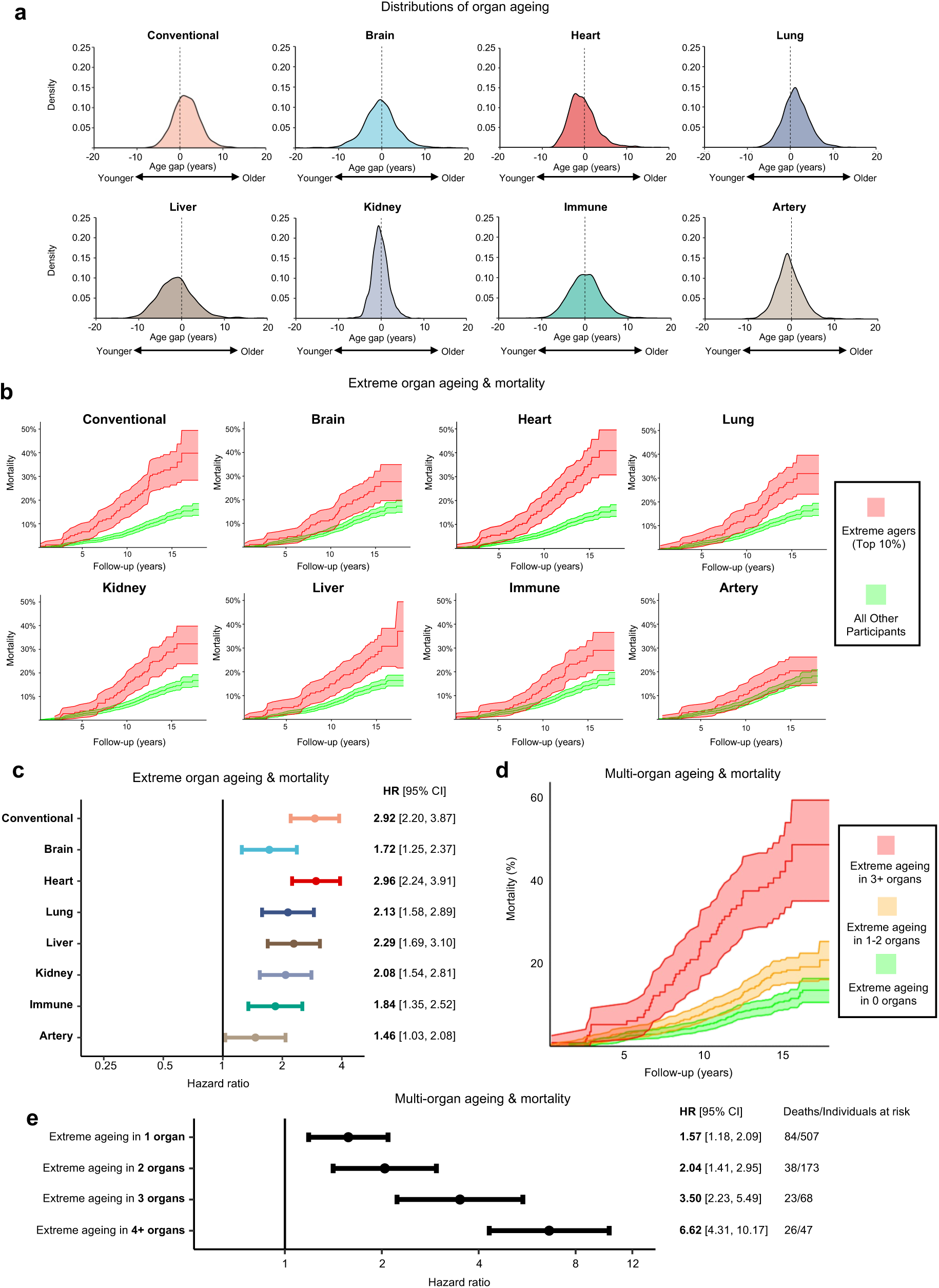
Organ ageing heterogeneity in a same-aged birth cohort holds prognostic value for mortality risk. **a**, Proteomic age gap distributions for 1,803 NSHD participants in late midlife. Age gaps reflect the participants’ estimated age relative to their chronological age: positive values indicate an older biological ageing phenotype, whilst negative values indicate a biologically younger phenotype. **b**, Kaplan-Meier estimates for cumulative incidence of death, demonstrating the relationship between extreme ageing and mortality risk. ‘Extreme agers’ were the oldest decile for each age gap distribution. **c**, Results from Cox proportional hazards models controlled for sex and chronological age for the Conventional age and each organ studied, comparing mortality risk for extreme agers to other participants. **d**, Kaplan-Meier estimates for cumulative incidence of death and **e,** Cox proportional hazards models demonstrating progressively increased mortality risk for individuals with multiple extremely aged organs versus those without any extremely aged organs.

We found that extreme ageing was linked to substantially elevated mortality risk **(Fig. 2b)**. Extreme ageing on the Conventional clock conferred 2.92 times the hazard (95% CI: 2.20-3.87, *p* = 1.05×10^-13^) of being amongst the 281 (15.6%) who died during follow-up **(Fig. 2c; Supplementary Table 6)**. Extreme ageing of each individual organ linked to greater mortality risk **(Fig. 2c)**, with extreme heart agers having the poorest prognosis (HR 2.96, 95% CI: 2.24-3.91, *p* = 2.72×10^-14^), consistent with cardiovascular disease as a dominant cause of premature death^24^. Individuals with extreme ageing in multiple organs (“multi-organ extreme agers”) were at especially increased risk **(Fig. 2d)**, with rising mortality for those with one (HR 1.57, 95% CI: 1.18-2.09), two (HR 2.04, 95% CI: 1.41-2.95), three (HR 3.50, 95% CI: 2.23-5.49) or four (HR 6.62, 95% CI: 4.31-10.17) extremely aged organs **(Fig. 2e; Supplementary Table 7)**.

### Life course factors shape organ ageing profiles

Tracking the considerable differences in the cohort’s life circumstances and behaviours for six decades after birth **(Extended Data Table 1; Supplementary Table 1)**, we examined whether these factors might shape biological ageing profiles, through linear regression adjusted for sex, chronological age and socio-economic status.

Several life course exposures associated plausibly with participants’ overall ageing profile, as measured by the Conventional clock **(Fig. 3)**. Being overweight in adolescence prospectively associated with accelerated ageing five decades later (*β* = 1.13 yrs, adj. *p* = 1.4×10^-4^; **Fig. 3a; Supplementary Table 8**), an association which persisted after adjusting for overweight in earlier-, mid- and late-adulthood **(Fig. 4a)**, implying that achieving healthy weight later in life may not mitigate the impact of being overweight in early life.

**Figure 3.**
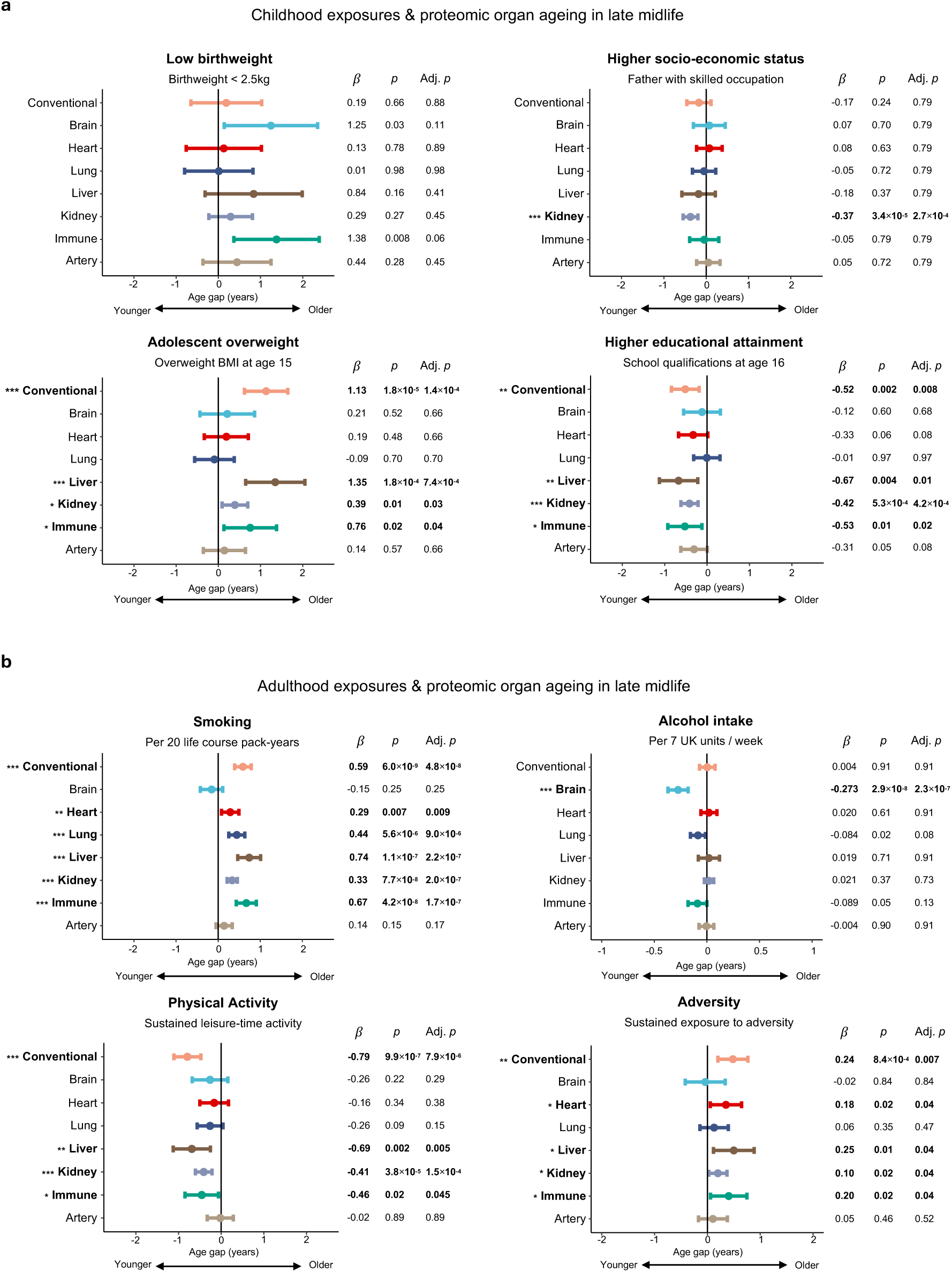
Life course exposures are connected to proteomic organ ageing profiles. **a**, Relationships between childhood exposures and Conventional or organ age gap examined with linear regression. *β* represents regression coefficient estimating the effect of the exposure on proteomic age gap, in years. Adjusted p value (adj. *p*) indicates *p* value after multiple comparisons correction using the Benjamini-Hochberg method. **b**, Relationships between exposures prospectively measured across adulthood and Conventional or organ age gap examined with linear regression. *β* represents regression coefficient and ‘adj. *p*’ indicates the *p* value after multiple comparisons correction.

**Figure 4.**
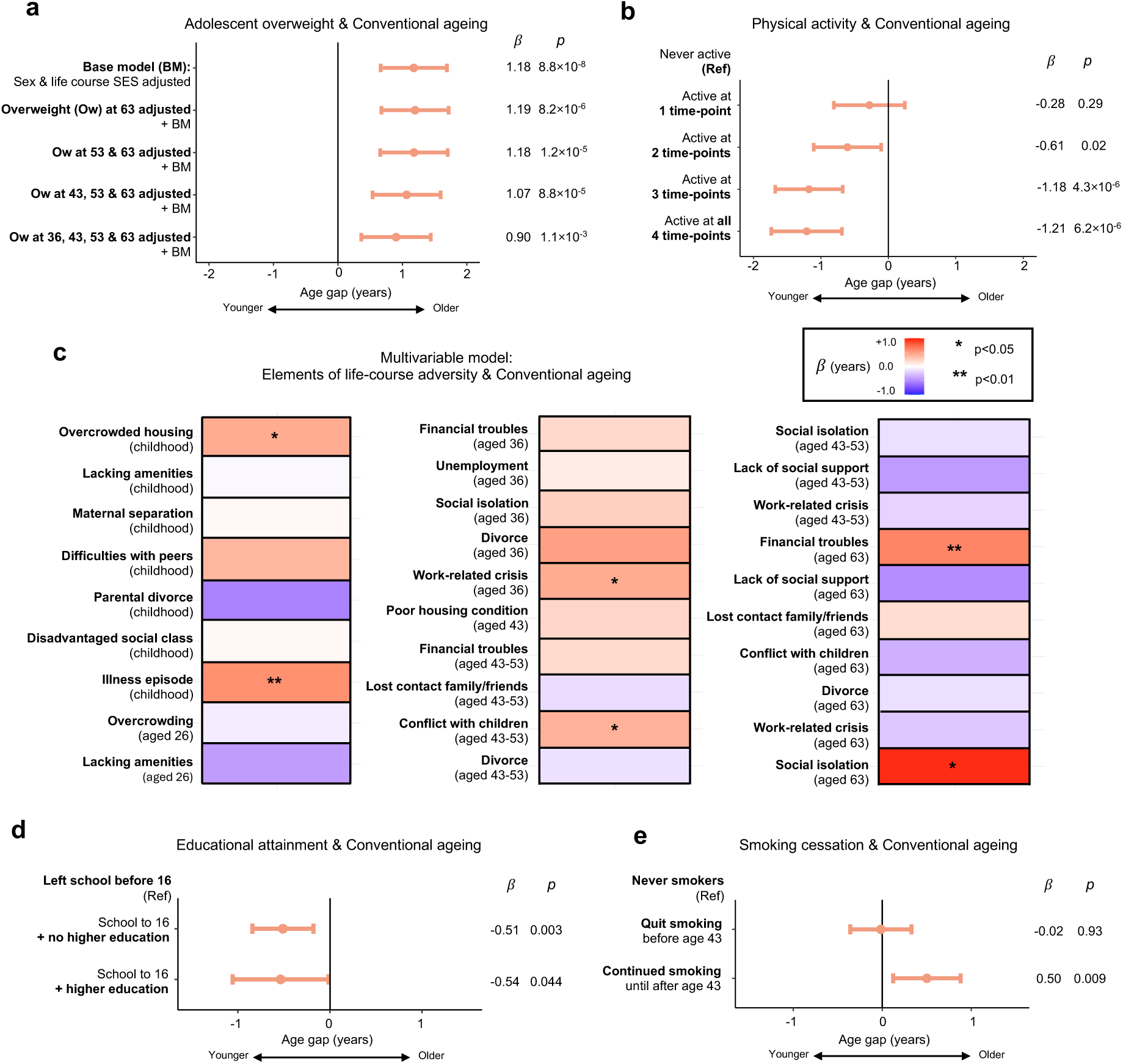
Life course analysis reveals cumulative and time-sensitive associations shaping proteomic ageing. **a**, Forest plot displaying linear regression analysis results of the relationship between overweight in adolescence and Conventional proteomic ageing. The association was robust to sensitivity analyses adjusting for overweight in earlier-, mid- and later adulthood. *ß* represents the regression coefficient for the effect of the exposure on organ age gap, in years, and *p* the p-value for each model. **b**, Forest plot displaying linear regression analysis results, revealing a dose-dependent relationship between physical activity levels across adulthood and Conventional proteomic ageing. **c**, Heatmap displaying results from multivariable linear regression analysis of the relationship between components of the life course adversity composite and Conventional proteomic ageing. Colour represents a positive (red, older) or negative (blue, younger) regression coefficient for Conventional age gap values in years. Significant results, where *p* < 0.05, are highlighted with an asterisk. **d,** Forest plot displaying linear regression results of the relationship between educational attainment groups and Conventional proteomic ageing. **e**, Forest plot displaying linear regression results of the relationship between stopping or continuing smoking in mid-adulthood and Conventional proteomic ageing, relative to never smokers.

Individuals completing school to age 16 were biologically younger in late midlife than their peers who did not (*β* = -0.52 yrs, adj. *p* = 0.008; **Fig. 3a; Supplementary Table 8**). The protective association of schooling was equivalently strong for individuals with or without a university degree **(Fig. 4d)**, suggesting that sustaining secondary education, rather than subsequently accessing higher education, might be the influential ingredient.

In adulthood, physical activity associated with younger age estimates (*β* = -0.79 yrs, adj. *p* = 7.9×10^-6^; **Fig. 3b; Supplementary Table 8**). Sustaining activity across adulthood was linked to a more advantageous ageing profile in a dose-dependent fashion **(Fig. 4b)**. Smoking strongly associated with older age estimates (*β* = 0.59 yrs per 20 pack-years, adj. *p* = 4.8×10^-8^; **Fig. 3b; Supplementary Table 8**). However, smokers who ceased by age 43 had no evidence of excess ageing relative to never smokers **(Fig. 4e)**, suggesting cessation by early mid-adulthood could mitigate impact.

Accumulating adversity across the life course was associated with accelerated ageing (*β* = 0.24 yrs per life period with adversity, adj. *p* = 0.007; **Fig. 3b; Supplementary Table 8**). Interrogating this composite through a multivariable model revealed that living in an overcrowded home (*β* = 0.42 yrs, *p* = 0.03; **Fig. 4c**) or suffering an illness episode (*β* = 0.57 yrs, *p* = 0.007) in childhood, experiencing conflict with children (*β* = 0.40 yrs, *p* = 0.02) or a work-related crisis (*β* = 0.43 yrs, *p* = 0.04) in mid-adulthood, and social isolation (*β* = 0.94 yrs, *p* = 0.03) or financial troubles (*β* = 0.61 yrs, *p* = 0.007) in later life independently associated with accelerated ageing, potentially reflecting different patterns of vulnerability to adverse experiences in each life period.

Investigating the determinants of organ-specific ageing, we found that liver, immune and kidney ageing associated with the same factors as the Conventional clock: adolescent overweight, smoking and cumulative adversity were linked to accelerated ageing, whereas lengthier school education and keeping physically active had protective associations (**Fig. 3)**.

We conducted exploratory protein-level analysis to investigate possible biological mechanisms underpinning these associations, discovering connections between adolescent overweight and the insulin-like growth factor system, which regulates insulin sensitivity **(Extended Data Fig. 3a; Supplementary Table 9)**; smoking and reduced longevity protein klotho and elevated renin **(Extended Data Fig. 3b; Supplementary Table 10)**; and physical activity and heme oxygenase (HMOX1) **(Extended Data Fig. 3c; Supplementary Table 11)**, a protein established to rise in response to exertion^25,26^.

Higher alcohol intake was associated with a younger brain age estimate (*β* = -0.27 yrs per 7 UK alcohol units per week, adj. *p* = 2.35×10^-7^; **Fig. 3b; Supplementary Table 8**). This relationship persisted after controlling for numerous confounders, including socio-economic status, educational attainment, social support, body habitus and physical activity **(Extended Data Fig. 4a)**. Relationships were robust over separate measures of alcohol intake taken at ages 36, 43, 53, and 63, and showed a dose-dependent pattern **(Extended Data Fig. 4b)**. Interrogation of the 201 brain-enriched proteins which comprise the brain ageing clock showed 21 targets (10.0%) were associated with alcohol intake at two or more time points, implying consistent biological signal **(Extended Data Fig. 4c; Supplementary Table 12)**. Feature importance for biological ageing (FIBA) analysis, which also accounts for each protein’s influence in determining the overall brain age estimate, determined a subgroup of proteins, including IgLON5, complexin-1 (CPLX-1), complexin-2 (CPLX-2) and neurexin-3 (NRXN-3), as particularly influential in driving the observed association **(Extended Data Fig. 4d; Supplementary Table 13)**.

### Maximising protective factors across life links to minimal extreme ageing risk

Life course exposures likely synergistically influence ageing rather than acting independently^13^. We hypothesised that an optimal combination of normal birthweight, higher socio-economic status, normal body mass index (BMI) and educational attainment in childhood, along with non-smoking, moderate alcohol consumption, sustained physical activity and lack of adversity across adulthood, would be maximally protective against accelerated ageing and mortality **(Fig. 5a)**. We calculated a protective factor score (PFS), summing how many protective factors cohort members had accrued across their life course. Participants split into five almost equal groups based on having ≤ 3 (19.2%), 4 (19.7%), 5 (23.3%), 6 (20.8%) or ≥ 7 (16.9%) of these protective factors **(Fig. 5b; Supplementary Table 14)**.

**Figure 5.**
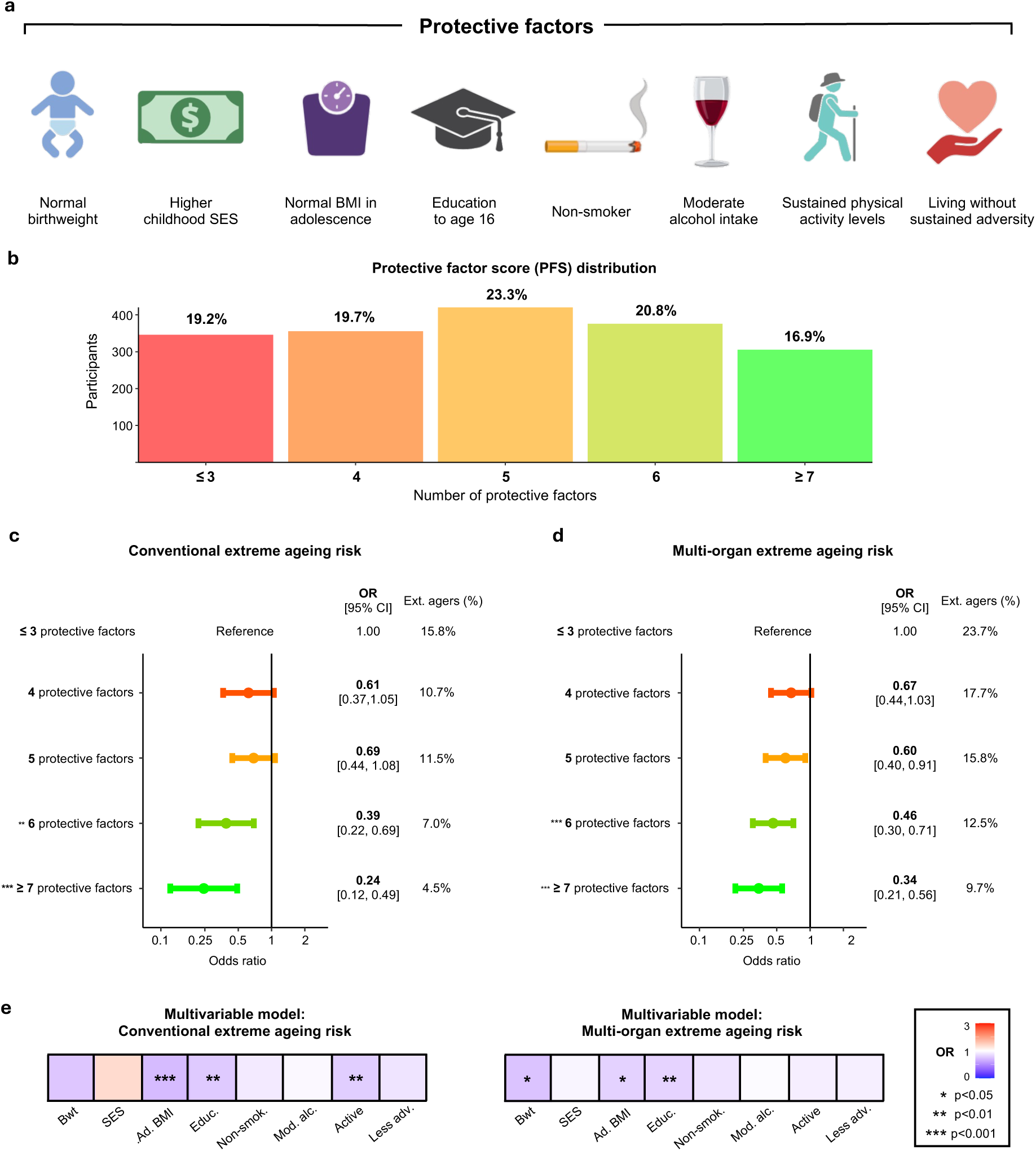
Accumulating protective factors across the life course associates with minimal extreme ageing risk. **a,** The eight life course factors we hypothesised to be protective for healthy ageing and longevity. Figure elements created with BioRender.com. **b**, Distribution of participants’ protective factor score (PFS): the number of protective factors they had accumulated across their life course. **c**, Logistic regression examined whether participants with more protective factors (higher PFS) had lower odds of extreme Conventional ageing. **d,** Logistic regression examined whether participants with higher PFS had lower odds of having extreme ageing in multiple organs (multi-organ extreme ageing). **e**, Multivariable models investigated which protective factors were associated with reduced odds of Conventional and multi-organ extreme ageing with mutual adjustment of other protective factors.

Higher PFS robustly associated with reduced odds of death by the end of follow-up (OR = 0.82 per unit increase in PFS, *p* = 1.5×10^-5^). Hypothesising that protective exposures promote longevity partly by reducing the risk of extreme ageing, we found that cohort members with the most protective life course exposures had four-fold lower odds of extreme Conventional ageing (OR = 0.24, 95% CI: 0.12-0.49, *p* = 8.1×10^-5^; **Fig. 5c; Supplementary Table 15**), and three-fold lower odds of having extreme ageing in multiple organs (OR = 0.34, 95% CI: 0.21-0.56, *p* = 1.9×10^-5^; **Fig. 5d; Supplementary Table 15**).

We used multivariable models to isolate the potential contribution of each protective factor, finding that childhood characteristics emerged as the dominant associations. Normal birthweight, healthy adolescent weight and sustained secondary education were all independently associated with reduced odds of having extreme ageing in multiple organs by late midlife **(Fig. 5e)**. Healthy adolescent weight and sustained education were also independently connected to lower odds of extreme Conventional ageing, as was maintaining activity across adulthood **(Fig. 5e)**.

Finally, we tested the overarching hypothesis: that favourable life course exposure profiles reduce odds of early death by influencing proteomic organ ageing. Mediation analyses adjusted for chronological age and sex estimated that differences in kidney ageing accounted for 22.9% (ACME = -0.008, *p* = 1.8×10^-5^, PM 95% CI: 6.8-39.0%**; Supplementary Table 16**) of the relationship between the PFS and mortality, with liver ageing contributing 11.7% (ACME = - 0.004, *p* = 0.002, PM 95% CI: 1.2-22.2%) and immune ageing 9.1% (ACME estimate = -0.003, *p* = 0.006, PM 95% CI: 1.2-17.0%), with no clear mediation via heart, brain, lung and artery ageing.

### Proteomic discovery identifies understudied longevity biomarkers

Taking advantage of 11k SomaScan (v5.0) data on 10,776 human plasma protein targets, we explored which proteins reflected an individual’s life course exposures and mortality risk in a single measure. Using differential abundance analyses corrected for multiple comparisons (Benjamini-Hochberg, FDR < 0.05), we determined that 3,328 proteins were associated with PFS **(Fig. 6a; Supplementary Table 17)**, 1,388 were associated with mortality **(Fig. 6b; Supplementary Table 18)** and 991 were linked to both **(Fig. 6c)**, underscoring the plasma proteome’s rich complexity and particular sensitivity to socio-behavioural exposures.

**Figure 6.**
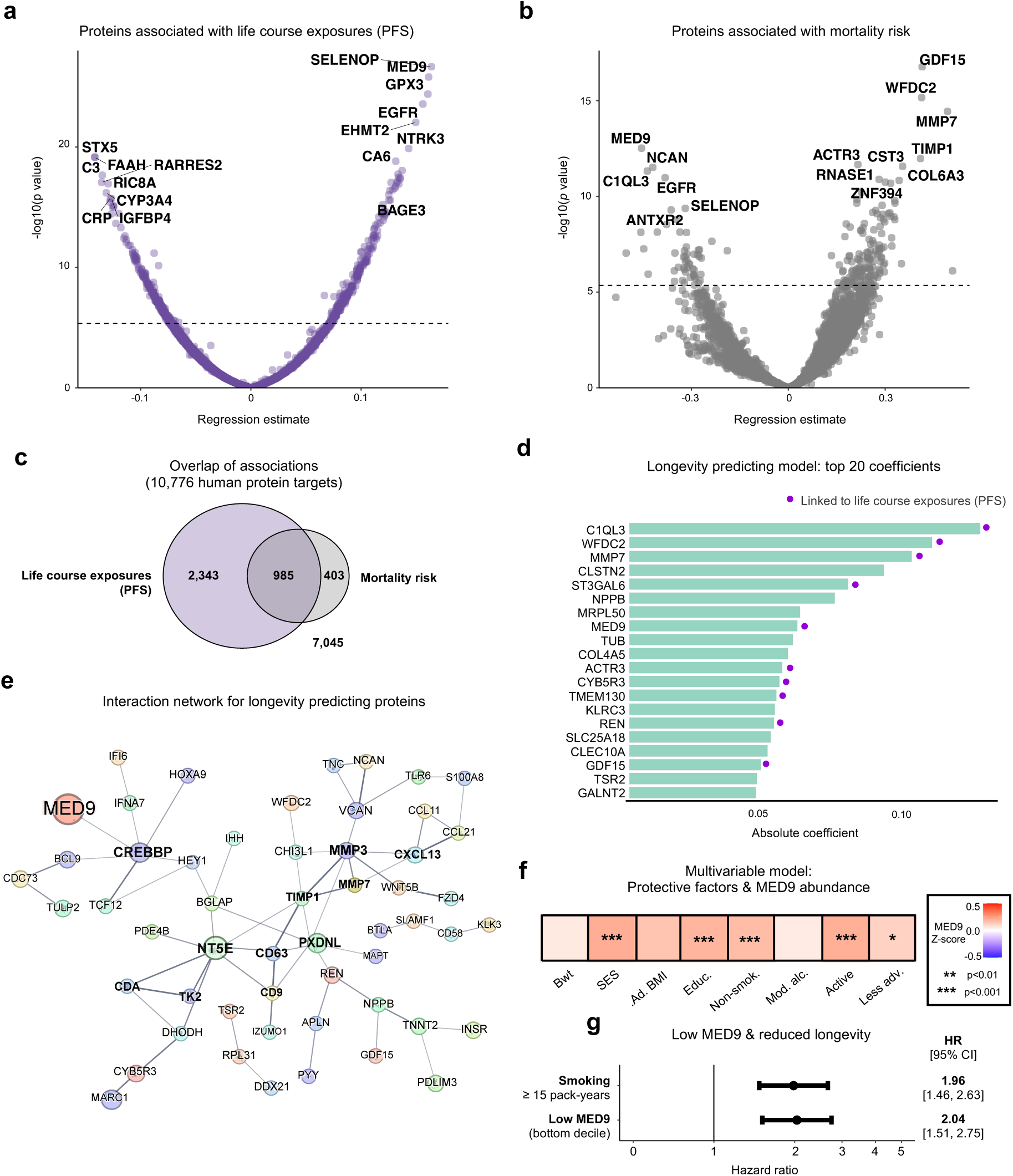
Proteins capturing life course exposures and mortality risk. **a**, Volcano plot of differential abundance analysis assessing the relationship between abundance of each of the 10,776 human protein targets detectable on the 11k SomaScan assay (v5.0) and the protective factor score (PFS). **b**, Volcano plot of results from Cox proportional hazards models analysing associations between the 10,776 protein targets and mortality. **c**, Venn diagram displaying the number of proteins associated with PFS and mortality after Benjamini-Hochberg (BH) multiple comparisons correction (FDR < 0.05). **d**, Bar graph showing the top 20 absolute coefficients in the elastic net-penalized Cox model predictive of longevity. Proteins significantly associated with the PFS (*p* < 0.05 after BH multiple comparisons correction) are highlighted with a purple dot. **e**, Interaction network of the proteins predictive of longevity, generated using the STRING website interface. Only proteins which exhibited interaction are visualised. **f**, A multivariable model indicated higher MED9 abundance was independently associated with higher socio-economic status (SES), educational attainment, non-smoking, activity and lack of adversity. **g**, Participants with low MED9 (bottom decile for protein abundance) were at increased mortality risk. The effect size was equivalent to the elevated mortality risk of smokers with 15 or more pack-years compared with never smokers.

We next used an elastic net-penalized Cox model to identify which combination of the 10,776 proteins had highest predictive value for longevity. A model of 143 proteins predicted mortality (Concordance Index: 0.70; top 20 shown in **Fig. 6d; Supplementary Table 19**), with many concurrently associated with the PFS, and predicted to engage in direct biological interaction (**Fig**. **6e**). The model included several widely established markers of health and disease, including Growth Differentiation Factor-15 (GDF-15), N-terminal pro-B-type natriuretic peptide (NT-proBNP), Troponin-T (TnT), Prostate-specific antigen (PSA) and Insulin receptor (INSR). However, nearly a third of the model’s proteins (42/143, 29.4%) are understudied in humans, not covered by previous SomaScan or Olink platforms **(Extended Data Fig. 5a)**.

Using Gene Ontology mapping to link this sub-group to diverse biological processes **(Extended Data Fig. 5b; Supplementary Table 20)**, we identified Mediator Complex Subunit 9 (MED9) as a candidate of particular interest. Of all 10,776 proteins examined, MED9 was the second most robustly associated with life course exposures (after SELENOP; adj. *p =* 8.2×10^-23^) and the eighth most influential in predicting longevity **(Fig. 6d; Supplementary Table 19)**. Higher childhood socio-economic status, educational attainment, non-smoking, physical activity and lack of adversity were independently associated with increased MED9 **(Fig. 6f; Supplementary Table 21)**. Conversely, participants with low MED9 (lowest decile for protein abundance) had double the odds of death, an effect comparable to smoking fifteen or more pack-years of cigarettes **(Fig. 6g; Supplementary Table 21)**. Given Mediator complex’s role as a master transcription regulator^27^, its subunit 9 (MED9) warrants further investigation as a potential biomarker for human longevity and therapeutic target.

## DISCUSSION

We demonstrate striking organ ageing heterogeneity amongst individuals with near-identical dates of birth, likely shaped by modifiable exposures across the life course. Results offer insights on which factors may influence how we age, when in life they might matter most, and how effects can biologically emerge, informing individual- or population-level strategies for healthy ageing promotion. Leveraging extensive proteomic coverage, we identified understudied proteins predictive of longevity, including MED9, which was independently related to a diverse range of life course socio-behavioural exposures.

Our study extends work using imaging^6,28^ or clinical^29^ phenotypes to demonstrate that organ ageing heterogeneity is present on the molecular level in closely comparable individuals of near-identical chronological age. Each organ’s trajectory was relatively asynchronous with others, consistent with, and potentially underpinning, differential susceptibility to organ-specific chronic disease^30^.

Accelerated ageing in all organs was associated with increased risk of subsequent death, with individuals having four or more organs with extreme ageing having more than six-fold risk of dying over the subsequent 15 years. This finding underscores the interdependence of bodily systems, suggesting that molecular loss of resilience across several organs confers a particular physiological vulnerability which is crucial to detect, since it may allow preventive efforts for averting early death to be initiated or focussed.

Life course analyses unearthed insights into the determinants of proteomic ageing in different life periods. Adolescent overweight was associated with accelerated ageing in later life, even when overweight in mid- and late-adulthood was controlled. This cohort was born in a time of food rationing, in stark contrast to the current environment in which almost a third of children are overweight^31^. Our findings suggest that those who are overweight as teenagers may still experience downstream adverse ageing effects even if they attain healthy weight in adult life, highlighting the need to tackle upstream obesogenic drivers^32^.

As expected, smoking associated with accelerated ageing in multiple organs. However, individuals who ceased smoking by 43 years were not more biologically aged than those who had never smoked, implying that promoting cessation at this early midlife turning point could yield long-term health rewards. Accumulating adversity across the life course was associated with accelerated proteomic ageing. Our data suggest that psychosocial and financial stressors, including living in overcrowded housing in childhood, family conflicts and work-related crises in midlife, and financial difficulties and social isolation in later life, can be reflected in molecular changes representative of health and longevity^33^. Leisure-time activity across adulthood was associated with reduced biological ageing in multiple organs, extending previous evidence that exercise is associated with superior cognitive performance^34^, and that even low-level exertion likely produces measurable benefit relative to sedentary lifestyle^35^. Individuals obtaining school qualifications at age 16 appeared biologically younger by late midlife than cohort members who had none, supporting the premise that completing secondary school education is a shielding influence for life course socio-economic trajectory with molecular correlates^36^.

Accumulating protective exposures from birth to late midlife was associated with a four-fold reduction in risk of extreme ageing. In a multivariable analysis, the protective associations of early-life exposures were most prominent, perhaps reflecting a crucial window for physiological development^37^, or that childhood circumstances set in motion chains of protection or risk which shape the remainder of the life course^13^.

Our findings suggest that kidney, liver and immune ageing are particularly sensitive to a broad range of socio-behavioural exposures. Examining the individual protein components of these ageing clocks indicates support for hypotheses on how life circumstances or behaviours become biologically embedded. Renin is the most influential protein in the kidney ageing clock, so ageing and up-regulation of the renin-angiotensin system, triggered by social or physiological stressors^38^, may explain its mediating influence. Imbalance of the immune ageing clock’s cytokines is a key mechanism driving chronic inflammation: an ageing hallmark^8^ linked to adverse socio-behavioural exposures^39^ and a range of pathophysiological processes. The insulin-like growth factor family of proteins are core components of the liver ageing clock, and their dysregulation is influential in driving impaired nutrient sensing^40^: an ageing hallmark^8^ associated with metabolic syndrome and reduced healthspan.

We observed that higher alcohol intake was associated with younger brain age in later life. Proteins involved in synaptic plasticity, neurotransmitter release and cellular adhesion, such as IgLON5 and complexin-1, were highlighted through interrogation, perhaps providing context on underlying molecular mechanisms. These findings require cautious interpretation. However, although alcohol excess clearly has many detrimental effects, previous investigation of the effect of moderate intake on brain function has produced mixed findings^41^. Beneficial associations between alcohol and cognition have been previously reported in the NSHD^42^ and other major cohorts^43–45^. We note that greater alcohol intake was recently associated with reduced ageing in other organs when proteomic ageing clocks were developed in the UK Biobank using Olink assays^46^. Our finding of an association between the brain-enriched plasma proteome and life course alcohol intake warrants experimental investigation to establish underpinning molecular mechanisms and the causative effects of moderate intake on the brain’s proteome, structure and function.

Among the 10,776 proteins assessed, 143 were selected as predictive of longevity, many of which were also associated with individuals’ life course socio-behavioural profiles. MED9 emerged as a compelling candidate, ranking among the top ten contributors to mortality prediction and second most robustly associated with protective exposures across life. MED9 forms part of the Mediator complex, a highly conserved molecular scaffold that orchestrates RNA polymerase II-driven transcription, acting as a central regulator of gene expression^27^. Its independent associations with early-life education and social class, physical activity, non-smoking, and lack of adversity suggests it may serve as a biomarker for modifiable factors influencing ageing trajectory. MED9 has not been widely studied in relation to human longevity and these data highlight it as a promising candidate for further mechanistic investigation and evaluation as a possible therapeutic target.

This study is not without limitations. Although the NSHD cohort provides unparalleled longitudinal depth, it lacks ethnic diversity, limiting generalisability to broader global populations. The use of observational data also means that causality cannot be assigned; reverse causation and residual confounding remain plausible. Age-related proteomic signatures, although reproducible and previously validated^4,9^, may incompletely capture underlying biological processes, as they are derived from specific datasets and could partly reflect cohort-specific characteristics. The study’s strengths include its integration of molecular, behavioural and survival data across nearly eight decades in a single geographically representative population. The ability to investigate organ-specific ageing within individuals of identical chronological age offers a rare opportunity to study biological variation. The use of independently verified proteomic clocks and repeated measures of life course exposures strengthens the reliability of findings and provides a framework that future studies can build upon.

In summary, these data trace a molecular signature of life course exposures, demonstrating that modifiable factors are likely to influence ageing trajectories via alterations in the plasma proteome. Organ ageing emerges as a biologically meaningful, multidimensional phenomenon — with substantial inter- and intra-individual heterogeneity, likely shaped by decades of exposures, and prognostic of longevity. Diverse socio-behavioural exposures associated with ageing across organs and individuals, while proteomic discovery highlighted both established and novel components of longevity biology. These findings emphasise both childhood and adult experiences as foundational for healthy ageing, underscoring the potential of life course-informed strategies to delay physiological deterioration, and the translational potential of proteomic tools to drive benefits for population health and patient care.

## METHODS

### Ethical Approval

This research was approved by the National Research Ethics Service Committee London (REC reference 14/LO/1173). All participants provided written informed consent.

### Participants

The MRC National Survey of Health and Development is a population-based birth cohort of 5,362 individuals born in a single week in March 1946^15^ **(Supplementary Table 1)**. Of the 5,362 total birth cohort participants, 3,163 remained alive and enrolled in the study at mean age of 63.2 years, when blood draw for plasma proteomics was undertaken **(Extended Data Fig. 1)**. 1,803 attended one of five regional UK locations (London, Manchester, Birmingham, Cardiff and Edinburgh) to undergo peripheral venepuncture, had plasma sampled for large-scale proteomics, and were included in the study **(Supplementary Table 1)**.

### Proteomic organ age estimates

The 11k SomaScan (v5.0) assay was run on plasma samples, which uses slow off-rate modified DNA aptamers (SOMAmers) to bind target proteins with high specificity, to record abundance for 10,776 human protein targets in Relative Fluorescent Units (RFU)^16^. SomaLogic’s standard procedures for normalization, calibration and quality control were applied to all samples^4,9^. To control for batch effects during assay quantification, pooled reference and buffer standards are included on each plate. Signal intensities are normalized within and across plates using the median values of the reference standards, followed by normalization to a pooled reference sample using an adaptive maximum likelihood procedure. Samples flagged by SomaLogic as having signal intensities significantly deviating from the expected range were excluded from analysis (n=4). To facilitate the use of ageing clocks which were developed on the previous 7k SomaScan (v4.1) assay, v.5.0 → v.4.1 multiplication scaling factors provided by SomaLogic were applied to the raw v.5.0 abundance values before organ age estimates were generated using the *organage* package in Python 3.8, producing the Conventional and organ-specific age estimates for the brain, heart, lung, liver, kidneys, immune system and arteries^4^. Consistent with previous approaches^4,9,12,47^, ageing phenotypes were expressed as age gaps, reflecting the difference between a participant’s estimated *biological age* and their *chronological age* at the time of sampling. An age gap of +1 indicates that a participant is estimated as one year biologically *older* than their chronological age, whilst an age gap of -1 indicates the participant to be estimated as one year biologically *younger* than their chronological age. Consistent with emerging evidence that the top decile of proteomic ageing is linked to reduced healthspan, we defined the oldest decile in each age gap distribution as ‘extreme agers’^12^.

### Mortality data

Participants were followed to age 78.3 years. Mortality data was drawn from NHS England and the NHS Central Register via NHS Digital in June 2024. For individuals who died during follow-up, deaths were dated to month and year. Individuals who emigrated or withdrew from digital health data collection during follow-up were censored (n=2).

### Life course exposures

Participants’ life course exposures were recorded from birth until proteomic sampling at mean age 63.2 years. We characterised eight factors across childhood and adulthood hypothesised to associate with proteomic organ ageing, selected based upon existing evidence linking the social determinants of health and health behaviours to epigenetic ageing^18,19,48–50^, prioritising measures with highest data quality in NSHD^15^. Childhood factors were low birthweight, childhood socio-economic status, adolescent overweight and educational attainment.

Birthweight in kilograms was recorded in March 1946 at the time of birth by a midwife or health professional present at the birth on a paper form. Consistent with the WHO’s definition^51^, all participants with a birthweight of less than 2.5 kg were classified as having low birthweight.

Childhood socio-economic status was assessed from questionnaires completed by the participants’ parents in 1950 (aged 4), 1957 (aged 11) and 1961 (aged 15). Father’s occupation was coded according to the 1970 OPCS Classification of Occupations. Individuals with father’s occupation categorised as professional, intermediate, or skilled non-manual were classified as ‘higher childhood socio-economic status’, whilst those coded with father’s occupation categorised as skilled manual, partly skilled or unskilled were classified as ‘lower childhood socio-economic status’.

Adolescent body mass index (BMI) was calculated from measures of participants’ height (measured to the nearest inch) and weight (measured to the nearest quarter of a pound) taken at age 15. WHO child growth charts were used to classify whether participants had BMI above threshold for overweight (female BMI cut-off: 23.5, male BMI cut-off: 22.7).

Educational attainment was assessed as the highest level of education attained to age 43. Individuals completing school to age 16 and obtaining O level qualifications were classified as ‘higher educational attainment’, whilst those who did not were classified as ‘lower educational attainment’ Smoking was assessed through questionnaires completed by participants at age 20, 25, 31, 36, 43, 53 and 63. A sum gave participants’ cumulative life course exposure to smoking, recorded in pack-years.

Alcohol intake was assessed through diet diaries completed over five days aged 36, 43, 53 and 63. Participants’ average daily intake was calculated from responses, and multiplied by seven to reflect average UK units of alcohol per week.

Physical activity was assessed through completion of leisure-time physical activity questionnaires at ages 36, 43, 53 and 63, as previously described^34^. At each timepoint, participants’ activity was categorised as inactive, mild or moderate-high. We classified individuals who had mild or moderate-high activity levels at two or more of the four timepoints as having engaged in sustained physical activity in adulthood.

Cumulative exposure to adversity was assessed using a previously published life course adversity measure^22^. The metric incorporates psychosocial (childhood maternal separation, difficulties with peers, parental divorce in childhood, divorce in adulthood, disadvantaged social class, losing contact with friends or family, conflict with children, social isolation and lack of social support), financial (overcrowded housing, lacking essential household amenities, financial troubles, work-related crises and unemployment) and physical (prolonged illness episode) sources of adversity at different periods in the life course. We included all prospectively assessed psychosocial and financial sources of adversity but omitted physical adversity related to adulthood illness episodes to reduce chance of reverse causation in analyses with biological ageing. We classified individuals who experienced adversity in at least three of the four life periods studied (childhood: age 0-18; early adulthood: age 18-36; mid-adulthood age 36-53; later adulthood: age 53-63) as having sustained exposure to adversity.

We hypothesised normal birthweight, higher childhood socio-economic status, healthy adolescent BMI, higher educational attainment, non-smoking, moderate alcohol intake (between 7 and 21 units per week), sustained physical activity and living without sustained adversity to be protective for healthy ageing. A protective factor score (PFS) was calculated as the number of these protective factors that applied to a participant’s life course.

### Statistical analysis

To account for missing life course data, we performed multiple imputation in R (v4.3.0) using the *mice* package. Continuous variables were imputed using predictive mean matching, binary variables using logistic regression and multi-level categorical variables using Bayesian polytomous regression, with inclusion of the study’s outcomes and health-related variables hypothesised to be predictive of missingness. Thirty datasets were imputed with 50 iterations. No proteomic or mortality data was imputed.

Cox proportional hazards models controlled for sex and chronological age and unadjusted Kaplan-Meier survival curves assessed relationships between extreme ageing and all-cause mortality, censored for emigration.

For primary analyses examining associations between life course exposures and proteomic ageing **(Fig. 3; Supplementary Table 8)**, linear regression controlled for sex, chronological age and socio-economic status was performed (see **Supplementary Information** for full details on model covariates). Each life course exposure was considered a distinct hypothesis and family of tests, with Benjamini-Hochberg (BH) multiple comparisons correction (FDR < 0.05) applied across results for the eight proteomic organ ageing clocks assessed (see **Supplementary Information** for full details on multiple comparisons strategy).

A series of secondary analyses were performed to deepen understanding of the relationship between life course exposures and proteomic ageing (see **Supplementary Information** for rationale and full details on model covariates). Targeted hypothesis-driven linear regression analyses were performed to further assess the associations observed in primary analyses between Conventional ageing and adolescent overweight, educational attainment, smoking, physical activity and life course adversity. Given the observed sensitivity of liver, immune and kidney ageing to socio-behavioural exposures, exploratory protein-level investigation was conducted to shed light on potential biological mechanisms. We conducted further analysis on the association between alcohol intake and reduced brain ageing to assess the robustness to potential residual confounding and biological plausibility of this finding. This included feature importance for biological ageing (FIBA) analysis, a previously described adaption of permutation feature importance, which aims to estimate of the relative contribution of each organ-specific protein in explaining observed associations^4^. Briefly, each single brain-specific protein is randomised in turn, brain age gap estimates are recalculated, and associations between brain age and alcohol intake are re-evaluated, noting each protein’s resulting ‘FIBA coefficient’: the difference between the original brain age-alcohol association coefficient and the observed coefficient after protein randomisation. The final FIBA coefficient for each protein is the mean FIBA coefficient across five randomisations.

Logistic regression adjusted for sex and chronological age examined the relationship between PFS and premature death or extreme ageing risk. Causal mediation analysis adjusted for sex and chronological age was performed with the *mediation* package in R (v4.3.0) separately for each organ-specific ageing clock to assess for its role as a mediator of the association between PFS and premature death.

Differential abundance analyses were performed across the 10,776 human proteins detected by the 11k SomaScan (v5.0) assay to examine associations between log10-transformed, standardized protein abundance and PFS with linear regression or mortality with Cox proportional hazards models. BH correction for multiple comparisons was applied. An elastic net-penalized Cox model with log link was applied to select which combination of the 10,776 proteins had the highest predictive value for longevity, performed with the *glmnet* package in R (v4.3.0). Model performance was measured using ten-fold cross-validated concordance index (C-index) and the regularization parameter λ was chosen by cross-validation at the value that maximized the C-index. The elastic net mixing parameter α was fixed at 0.5. STRING network analysis was used to illustrate interactions between proteins (nodes) based on known and predicted relationships (edges) in the <u>STRING database</u>. Proteins included in the network exceeded a minimum interaction score threshold of 0.5, taking into account all interaction data sources available in the STRING database. Proteins were annotated to Gene Ontology (GO) terms using *org.Hs.eg.db* in R (v4.3.0), restricted to biological processes ontology. Related GO terms were collapsed into broader functional categories (e.g., immune response, inflammatory response, cell adhesion), yielding a binary protein–category matrix for visualization and analysis.

## ACKNOWLEDGEMENTS

We thank the participants of the MRC National Survey of Health and Development (NSHD) for their longstanding contributions. We are grateful to the NSHD study team for support with data collection and study coordination. We thank A. Isakova, P. Moran-Losada and other members for feedback and A. Raghavan for assistance with data cleaning.

## AUTHOR CONTRIBUTIONS

J.W.G, J.M.S. and T.W.-C. conceptualized the study. J.W.G. led study design, analysis and visualisation. J.M.S. and T.W.-C. supervised the study. J.N provided statistical input. V.A.B. and D.Y.D. contributed to analysis and/or visualisation of protein-level data. A.F., H.S.O., S.-N. J., D.W., J.K.R. and N.C. provided analytical input. A.W. co-ordinated resources, including sample handling. J.W.G. produced figures and wrote the manuscript. J.M.S. edited the manuscript. All authors critically revised the manuscript for intellectual content. All authors read and approved the final version of the manuscript.

## DATA AVAILABILITY

Bona fide researchers can apply to access the NSHD data via a standard application procedure (further details available at https://skylark.ucl.ac.uk/NSHD/access/). Mortality data can be requested from the UK Longitudinal Linkage Collaboration (https://ukllc.ac.uk/).

## CODE AVAILABILITY

A GitHub repository containing analysis code can be found at: https://github.com/jwgroves/NSHD-proteomic-organ-ageing.

## ADDITIONAL INFORMATION

Supplementary Information is available for this paper. Correspondence and requests for materials should be addressed to James W Groves, Tony Wyss-Coray and Jonathan M Schott.

## EXTENDED DATA FIGURES & TABLE

**Extended Data Figure 1.**
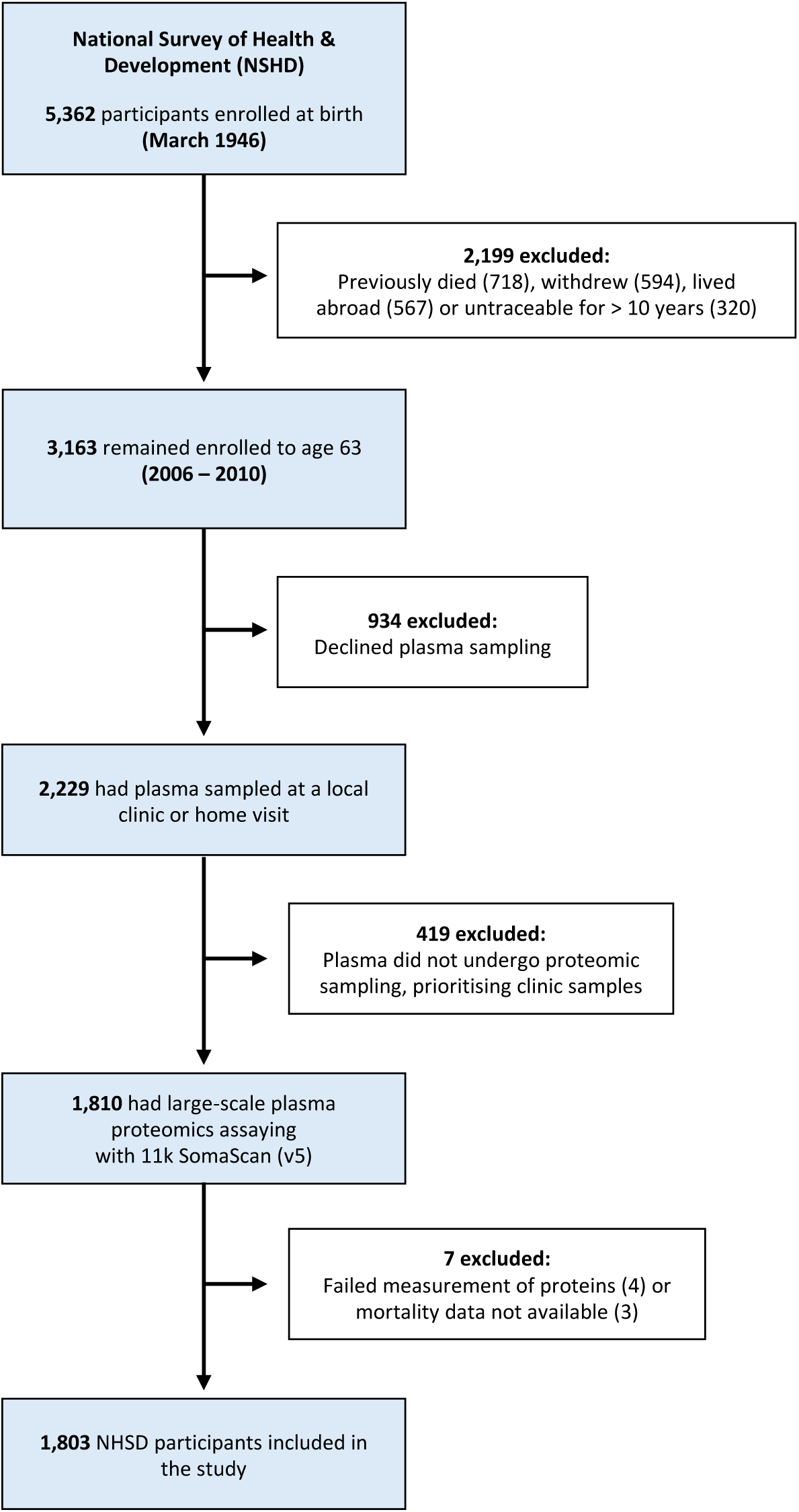
Study participants.

**Extended Data Figure 2.**
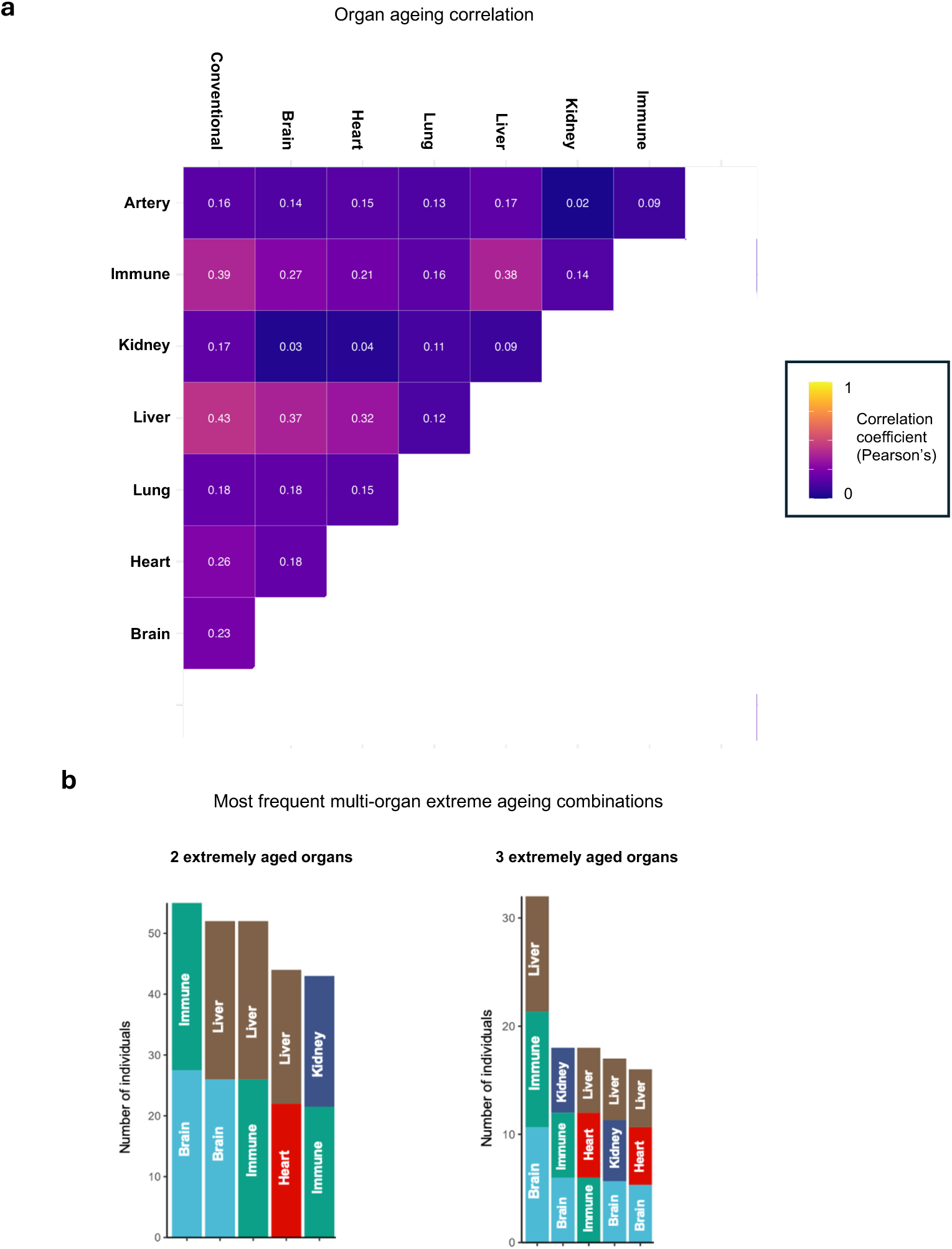
Heterogeneous patterns of organ ageing. **a**, Pearson’s correlation coefficient matrix for Conventional and organ-specific age gaps. **b**, Counts of the most frequent organ combinations showing concurrent extreme ageing. The top five combinations are shown for individuals with at least two or at least three extremely aged organs.

**Extended Data Table 1.** Participant demographics, life course exposures and mortality. Summary statistics for participants’ demographic information, life course exposures and mortality. Continuous variables are presented as mean (SD) and categorical variables as counts (%). For life course exposures, statistics were pooled across the thirty multiple imputation datasets.

| Variable | Category | n (%) \| Mean (SD) |
| --- | --- | --- |
| Demographic information |  |  |
| Age at proteomics (yrs) |  | 63.2 (1.1) |
| Sex | Female | 913 (50.6%) |
|  | Male | 890 (49.4%) |
| Ethnicity | White Caucasian | 1803 (100%) |
| Childhood life course exposures |  |  |
| Low birthweight (<2.5kg) | No | 1750.6 (97.1%) |
|  | Yes | 52.4 (2.9%) |
| Relative childhood social class<br>(paternal occupation) | Professional | 139.2 (7.7%) |
|  | Intermediate | 404.5 (22.4%) |
|  | Skilled non-manual | 378.8 (21.0%) |
|  | Skilled manual | 525.7 (29.2%) |
|  | Partly skilled | 309.7 (17.2%) |
|  | Unskilled | 105.1 (5.8%) |
| Educational attainment<br>(secondary education qualifications) | No | 653.9 (36.3%) |
|  | Yes | 1149.1 (63.7%) |
| Adolescent overweight<br>(BMI aged 15) | No | 1614.3 (89.5%) |
|  | Yes | 188.7 (10.5%) |
| Adulthood life course exposures |  |  |
| Smoking<br>(cumulative pack-years) |  | 9.9 (15.0) |
| Alcohol<br>(mean units per week) |  | 16.8 (15.0) |
| Physical activity in adulthood | Never active | 240.5 (13.3%) |
|  | Active at 1 of 4 time-points | 360.5 (20.0%) |
|  | Active at 2 of 4 time-points | 425.4 (23.6%) |
|  | Active at 3 of 4 time-points | 426.8 (23.7%) |
|  | Active all 4 time-points | 349.8 (19.4%) |
| Adversity across the life course<br>(financial/psychosocial) | No adversity in any life period | 37.8 (2.1%) |
|  | Adversity in 1 of 4 life periods | 266.9 (14.8%) |
|  | Adversity in 2 of 4 life periods | 514.7 (28.5%) |
|  | Adversity in 3 of 4 life periods | 640.9 (35.5%) |
|  | Adversity all 4 life periods | 342.8 (19.0%) |
| Longevity outcome |  |  |
| Mortality | Survived | 1522 (84.4%) |
|  | Died | 281 (15.6%) |

**Extended Data Figure 3.**
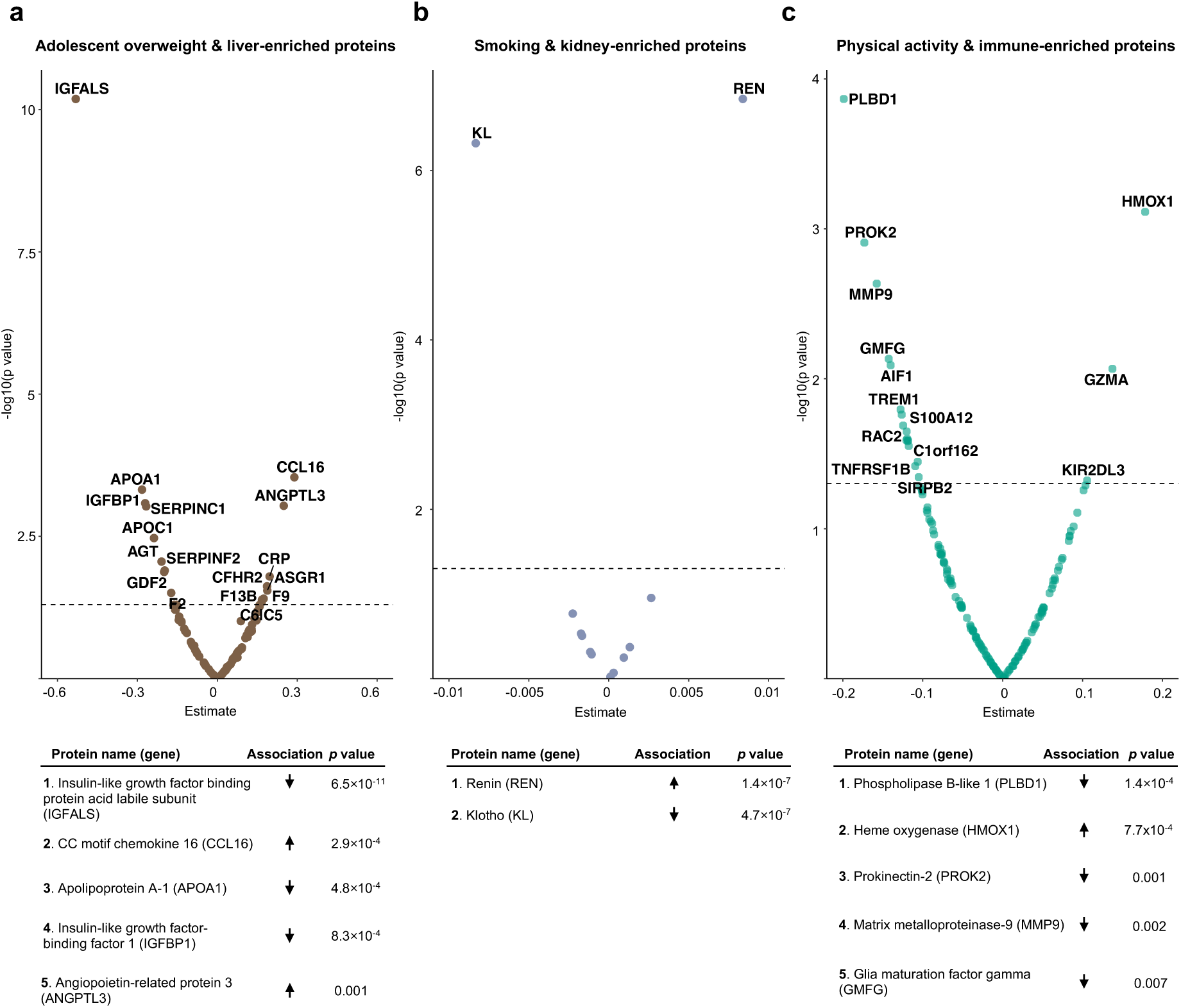
Exploring organ-specific proteomic correlates of life course exposures. **a**, Volcano plot of associations between liver-enriched proteins and adolescent overweight, with the most statistically significant hits summarised below. **b**, Volcano plot of associations between kidney-enriched proteins and smoking. **c**, Volcano plot of associations between immune-enriched proteins and activity.

**Extended Data Figure 4.**
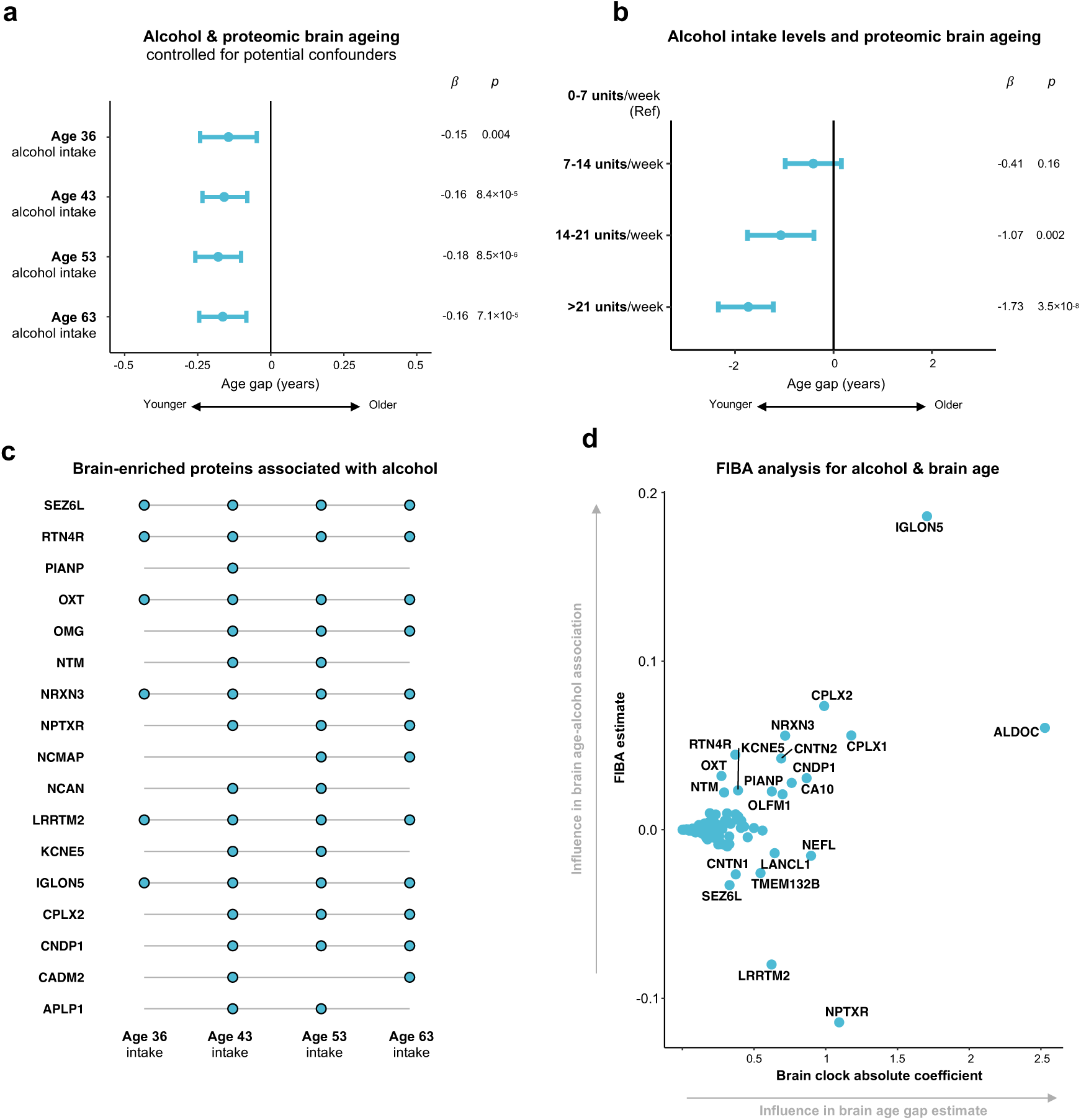
Scrutinising the association between alcohol intake and proteomic brain age. **a**, Linear regression results with brain age gap as the outcome and each separate measure of alcohol intake as the predictor (assessed at ages 36, 43, 53, 63), controlled for a range of confounders, including childhood and adulthood socio-economic status, educational attainment, social support, body habitus and physical activity. **b**, Linear regression results assessing for a dose-dependent relationship between alcohol intake and brain age gap. **c**, Brain-enriched proteins associated with alcohol intake measurements at two or more timepoints, assessed through linear regression for all 201 brain-enriched proteins with Benjamini-Hochberg multiple comparisons correction. **d**, Feature importance for biological ageing (FIBA) analysis to determine brain-enriched proteins most influential in driving associations between brain age and alcohol intake. As previously described^4^, the analysis randomises each brain-enriched protein in turn to estimate its impact on the association of interest (**see Methods**). A high FIBA estimate (y axis) indicates that a protein is estimated to hold an influential role in this association, based on the observed data. The brain ageing clock absolute coefficient (x axis) represents a protein’s influence in determining a participant’s brain age gap estimate.

**Extended Data Figure 5.**
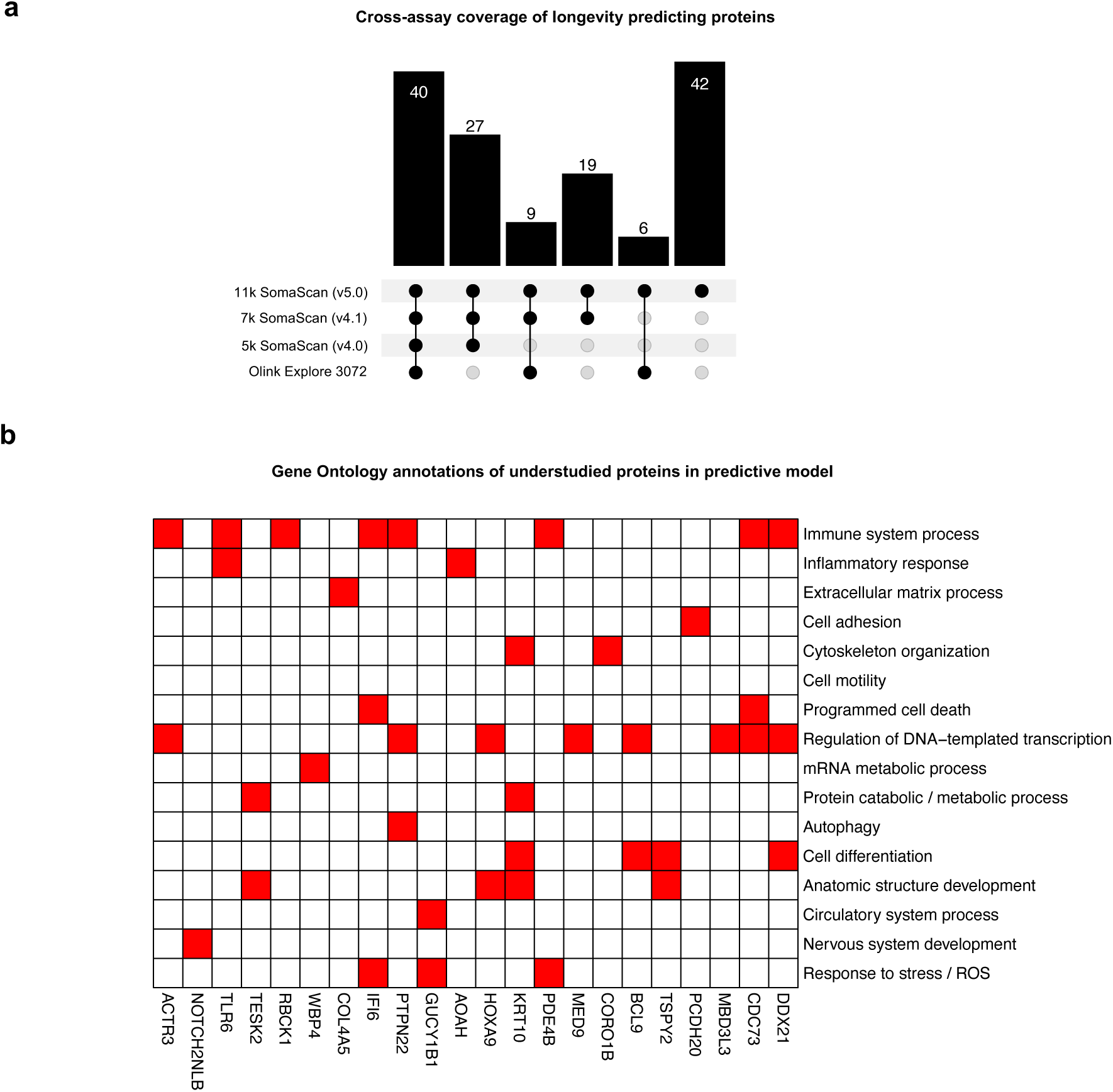
Understudied proteins in longevity predicting model. **a**, Upset plot summarising coverage of the 143 longevity-predicting proteins across widely used large-scale proteomic assays. 40 (28.0%) were common to all assays displayed. We took interest in the 42 understudied proteins not detectable on previous assays. **b**, Gene Ontology slim mapping annotations for understudied proteins predictive of longevity. Only proteins with annotation hits are shown.

